# The potential cost-effectiveness of controlling dengue in Indonesia using *wMel Wolbachia* released at scale: a modelling study

**DOI:** 10.1101/2020.01.11.20017186

**Authors:** Oliver J. Brady, Dinar D. Kharisma, Nandyan N. Wilastonegoro, Kathleen M. O’Reilly, Emilie Hendricx, Leonardo S. Bastos, Laith Yakob, Donald S. Shepard

## Abstract

**Background:** Release of virus-blocking *Wolbachia* infected mosquitoes is an emerging disease control strategy that aims to control dengue and other arboviral infections. Early entomological data and modelling analyses have suggested promising outcomes and *wMel Wolbachia* releases are now ongoing or planned in 12 countries. To help inform potential scale-up beyond single city releases, we assessed this technology’s cost-effectiveness under different programmatic options.

**Methods:** Using costing data from existing *Wolbachia* releases, previous estimates of *Wolbachia* effectiveness, and a spatially-explicit model of release and surveillance requirements, we predicted the costs and effectiveness of the on-going programme in Yogyakarta City and three new hypothetical programmes in Yogyakarta Special Autonomous Region, Jakarta and Bali.

**Results:** We predicted *Wolbachia* to be a highly cost-effective intervention when deployed in high density urban areas with gross cost-effectiveness ratios below $1,500 per DALY averted. When offsets from the health system and societal perspective were included, such programmes even became cost saving over 10-year time horizons with favourable benefit-cost ratios of 1.35 to 3.40. Sequencing *Wolbachia* releases over ten years could reduce programme costs by approximately 38% compared to simultaneous releases everywhere, but also delays the benefits. Even if unexpected challenges occurred during deployment, such as emergence of resistance in the medium-term or low effective coverage, *Wolbachia* would remain a cost saving intervention.

**Conclusions:** *Wolbachia* releases in high density urban areas is expected to be highly cost-effective and could potentially be the first cost saving intervention for dengue. Sites with strong public health infrastructure, fiscal capacity, and community support should be prioritized.

## Background

The mosquito species *Aedes aegypti* and *Ae. albopictus* are responsible for transmitting a range of growing global arboviral infections. Existing vector control tools alone have been unable to sustainably control these mosquito species or the diseases they transmit [1], and a range of novel technologies are under development [2].

One such novel intervention is release of mosquitoes infected with the intracellular bacterium *Wolbachia* [3]. Mosquitoes infected with *Wolbachia* are i) less likely to disseminate dengue, chikungunya, Zika and yellow fever viruses and thus are less likely to become infectious [3–5] and ii) can suppress or replace the natural mosquito population due to fatal cytoplasmic incompatibility among *Wolbachia*-wild type mating pairs [3]. *Wolbachia* can, therefore, be used to either replace the existing mosquito population with a lower competence phenotype by releasing females or suppress existing population by releasing males. To date, 13 countries have ongoing replacement programmes at various stages of development, with 12 through the World Mosquito Programme (WMP, www.worldmosquitoprogram.org) and an independent program in Malaysia [6]. Meanwhile China [7], Singapore [8], and the USA [9] have chosen to use suppression-based programmes due to perceived greater compatibility with their existing intensive and long term efforts to suppress mosquito populations.

Replacement programmes with *Wolbachia* entail substantial initial investments to establish *Wolbachia* in the mosquito population through intensive releases at the beginning of the programme but potentially offer considerable long-term benefits. The replacement approach contrasts with suppression strategies with *Wolbachia*, sterile insect techniques or conventional vector control tools, which likely need ongoing application. Both approaches are in the early stages of gathering entomological and epidemiological evidence of effectiveness [2]. Among these novel methods, replacement with *wMel Wolbachia* has, arguably, the most developed evidence base so far [10] because it has demonstrated replacement in multiple sites [11, 12], durability of unaltered replacement since 2011 in Townsville, Australia [13], reductions in reported dengue cases in observational study designs in five countries [14], and a cluster randomised trial is currently underway in Yogyakarta, Indonesia [15] with epidemiological outcome results expected in late 2020. Recent events including the 2015-7 Latin American Zika outbreak and the record breaking 2019 global dengue outbreak have hastened the adoption of novel *Aedes* control tools. Given the acute need to make decisions on adoption, mathematical models can be used to predict impact in different areas long before field data become available [16, 17]. Pairing these epidemiological predictions with intervention cost and cost-of-illness data enable cost-effectiveness calculations that can inform decisions on *Wolbachia* scale up.

One such priority setting is Indonesia, where city-wide *Wolbachia* releases are already planned in Yogyakarta City after the randomised trial [15]. In 2016 Indonesia launched its “Healthy Indonesia Program with Family Approach”, which includes cleaning the environment and addressing communicable diseases, including malaria and dengue [18]. This program provides encouragement and some national funding. In addition, in the Yogyakarta Special Autonomous Region (SAR), the governor has confirmed his support for novel technologies, including *Wolbachia* [19], suggesting support for expansion beyond Yogyakarta City.

Cost effectiveness analyses (CEAs) have proved instrumental for early adoption of a number of interventions, including for *Aedes*-borne pathogens. Fitzpatrick et al. estimated that, assuming they were 70-90% effective, conventional *Aedes* suppression tools would achieve cost-effectiveness ratios of between $679-$1331 per Disability-Adjusted Life Year (DALY) averted (2013 USD) [20]. The recently developed dengue vaccine, Dengvaxia®, also included model-predicted CEA as part of its feasibility assessment, with predictions without sero-testing of $11-44 per DALY averted (2014 USD) [21]. Dengvaxia® has also been estimated to be highly cost effective ($1,800 per DALY, health systems perspective) or cost saving ($-1,800 per DALY, societal perspective) under the WHO’s modified test-and-vaccinate recommendation, however with more limited overall impact (14.3% reduction in hospitalisation) [22].

Here we use existing *Wolbachia* release cost and programme data to build a model that predicts cost of release in different environments. We merge the cost predictions with previously published estimates of *Wolbachia* effectiveness [16] to assess cost-effectiveness and its sensitivity to different programmatic options for consideration at the next stage of scale up of this technology.

## Methods

### Phases of the programme

In this analysis we conceptually divide a potential *Wolbachia* replacement programme in a given city into four phases based on previous WMP operations. We do not consider the additional costs of obtaining regulatory approval in Indonesia as *Wolbachia* release has already been approved by the local Yogyakarta SAR government and the national government already has an active involvement in the project as part of the independent data monitoring committee. Phase 1 (“Setup”, 2 year duration) includes establishing insectaries, laboratories, site offices, local regulatory approval, hiring staff and planning and administering the programme and pre-release community engagement. Phase 2 (“Release”, 1 year duration) involves release of *Wolbachia* mosquitoes over target areas applying the resources established during Phase 1. In Phase 3 (“short-term monitoring”, 3 years duration), ongoing surveillance of the mosquito and human population is conducted in the release area. Phase 4 (“long-term monitoring”, 7 years duration) entails reduced entomologic monitoring as the intervention proves its reliability.

In the existing program in Yogyakarta City, programme setup took four years; however, this included gaining national approval, design of the cluster randomised trial, all Phase 1 activities and release in half the city. We anticipate faster setup times of subsequent programmes elsewhere in Indonesia due to the experiences and approvals gained in Yogyakarta City.

For the main analysis we consider two speed scenarios: i) an “accelerated” scenario, with every area conducting Phases 1-4 simultaneously and independently (total programme length 13 years), and ii) a “sequenced” scenario, in which Phase 2 releases are spread over 10 years with certain centralised resources moved or re-utilized across different locations (total programme length 20 years, Appendix page 7).

### Costing *Wolbachia* releases (Phases 1 and 2)

We hypothesised that the main determinants of the cost of releasing *Wolbachia* per square kilometre (km) were directly or indirectly related to: i) the human population density in the release area, ii) release material (adult or egg mosquitoes), iii) local cost of labour (as measured by country Gross Domestic Product adjusted for Purchasing Power Parity (GDP PPP)[23]) and iv) phase of the programme. Previous *Wolbachia* releases have shown that higher human density areas require higher mosquito release numbers per unit area because they typically have higher natural mosquito population sizes, hence raising costs [24]. Transportation costs of *Wolbachia*-infected mosquito eggs are lower than for adult releases because they can be distributed by the postal system and because the community can undertake releases; however, this can also increase community engagement costs. Adult releases require specific equipment and personnel to drive around the target area and conduct releases at pre-specified sites, but can potentially be achieved more quickly.

Data were extracted from WMP budgets for releases in Indonesia, Colombia, Sri Lanka, Australia and Vanuatu (Appendix page 1). These data were used to fit a generalised linear regression model between cost per km^2^ of release area and the above covariates (Appendix page 2).

#### Costing long-term surveillance (Phases 3 and 4)

Our estimates of the long-term monitoring costs of a *Wolbachia* programme (Phase 3 and 4) build on a detailed budget analysis developed by the WMP for Phase 2 of the programme in Yogyakarta City. This includes a gradual reduction in monitoring intensity with corresponding budget reductions (Appendix page 7).

### Cost of dengue illness averted

We obtain current fine-scale (5km x 5km) estimates of the current case burden of dengue in Indonesia and the economic cost per case in different treatment settings and from different payer perspectives from recent parallel studies. [16, 25] These maps of burden were downscaled to 100m x 100m spatial resolution to match the resolution of population datasets using bilinear interpolation.

### Candidate release sites

In this analysis we produce estimates for four candidate sites. (1) Yogyakarta City, (2) remaining areas in Yogyakarta SAR, (3) most of the special capital region of Jakarta (excluding Kepulauan Seribu [Thousand Islands] Regency), and (4) the island of Bali.

We chose this focus based on a combination of political, economic, and epidemiologic considerations. The Yogyakarta City trial carried an ethical and political expectation to assess expanding releases across the rest of Yogyakarta City and the remainder of the SAR. The other two candidate sites, Jakarta and Bali, are two of the country’s most important economic regions as commercial and tourism hubs. In epidemiologic terms, high density cities, such as Jakarta and Denpasar, Bali have a disproportionately high concentration of national dengue burden [16] and island-wide releases are likely to minimise the risk of re-introduction of native *Ae. aegypti* populations.

Within each of these sites, not every area is expected to be covered by *Wolbachia*. We consider only areas with a human population density of at least 1,000 people per km^2^ as eligible for *Wolbachia* releases. Previous WMP releases in Townsville and Cairns, Australia have proven the ability to establish *Wolbachia* in areas approaching 1,000 people per km^2^ (Figure 1), but based on existing programme experience, lower population densities are likely to prove prohibitive to natural mosquito dispersal and may significantly increase the cost or lower persistence of *Wolbachia* mosquitoes.

**Figure 1:**
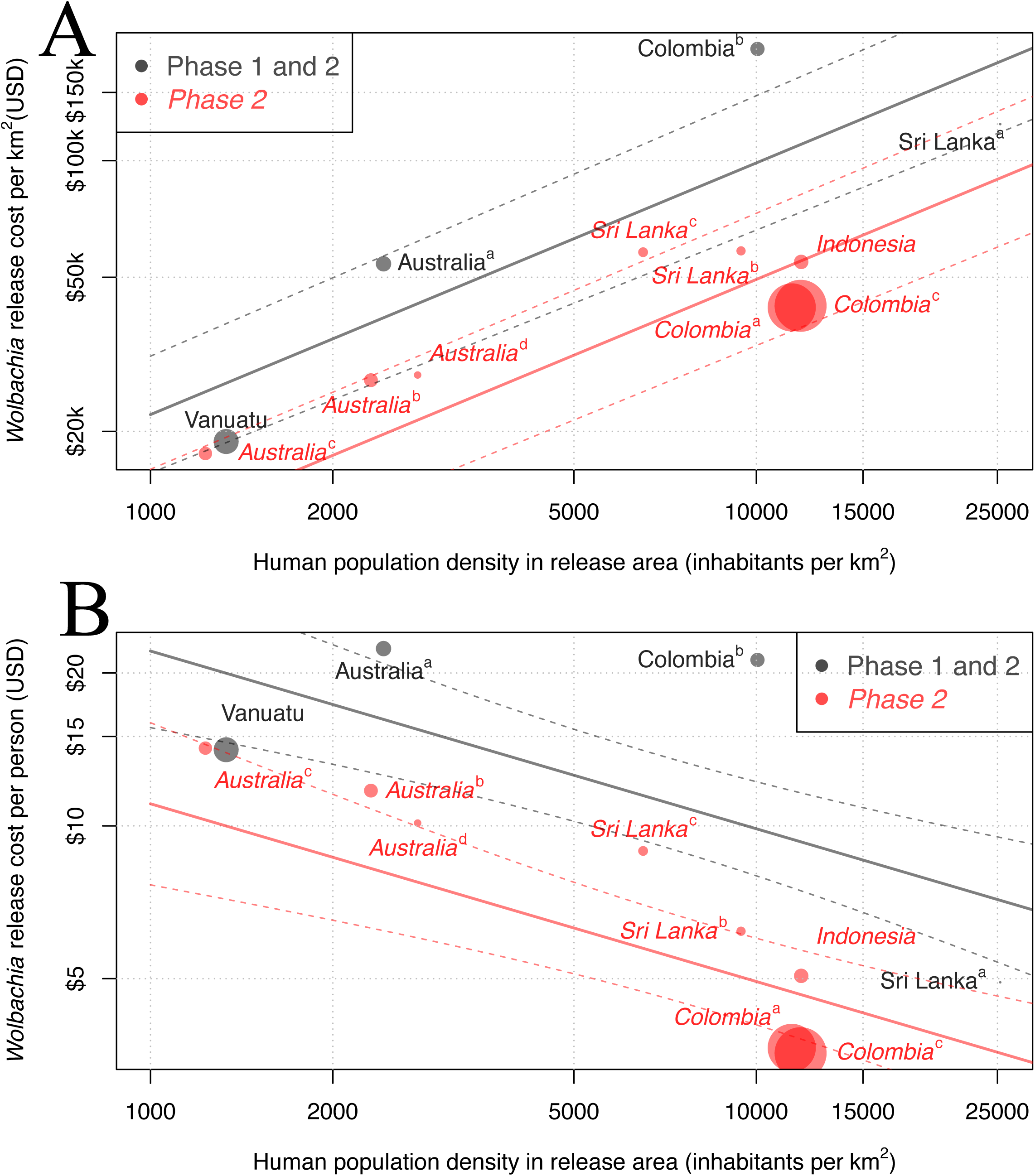
The fitted relationship between human population density and projected cost of deployment of Wolbachia per km^2^ (A) and cost per person (B). All axes are on log_10_ scales. The cost per km^2^ model fit mean (solid lines) and standard error (dashed lines) for each programme phase are shown. Circle area is proportional to size of release area in each site.

#### Time horizon, acquisition of benefits and discounting

As *Wolbachia* is an early stage technology, we take a conservative approach to our calculation of cost-effectiveness. We only assume benefits of *Wolbachia* persist for 10 years post completion of releases in the target area (i.e. benefits only accumulated in Phases 3-4, Appendix page 7) based on the duration of continued persistence of *Wolbachia* releases in northern Queensland since 2011 [26].

We assumed that the number of cases averted would be the same each year. All costs and benefits were discounted at 3% per annum [27], calculated at the end of each year. The cost of the programme was predicted using population data at 100m x 100m resolution from Worldpop [28] assuming a programme using egg releases with Indonesia’s national-2018 per capita GDP (PPP, $12,378).

Two measures of cost-effectiveness are shown. First, from a health systems perspective (gross cost-effectiveness), where the investment cost of the programme is divided by the number of DALYs averted over the 10 years post deployment (discounted at 3% per year). Second, from a societal perspective (net cost-effectiveness), where offsets to direct medical treatment costs are first deducted from the programme investment costs. Benefit-cost ratios are also calculated from health systems and societal perspectives separately.

#### Sensitivity analyses

To assess the sensitivity of our predictions to uncertainties in various inputs to our model we performed a univariate sensitivity analysis based on the 2.5% and 97.5% estimates for each of the following parameters: i) case burden, ii) *Wolbachia* effectiveness, iii) cost of *Wolbachia* releases, and iv) cost per episode of dengue illness. In addition, we also examined the sensitivity of cost-effectiveness to several hypothesised environmental and genetic challenges and changes that may occur as a consequence of *Wolbachia* introduction [29]. These include i) low coverage (50% vs baseline 100% coverage), ii) releases that are initially uncompetitive with wild-type mosquito populations and iii) emergence of resistance (after 5 years). The cost-effectiveness of programme modifications to address these challenges are also assessed. Furthermore, we predict the cost-effectiveness of future cost-saving adaptations of the program including iv) reliance on passive disease surveillance (as opposed to continued entomological surveillance in Phase 4 of the programme) and v) generic innovations, efficiencies and economies of scale that reduce the cost base of the programme by 50%. Further details on rationale for these scenarios and their parameterisation are available in the Appendix page 3. All analyses were performed in R version 3.6.1. with all code publicly available in the following GitHub repository (https://github.com/obrady/Wolbachia_CE/tree/V1).

## Results

### *Wolbachia* programme costs

The results of our model to predict the cost of releasing *Wolbachia* mosquitoes in new areas using existing programme budgets is shown in Figure 1. This model identified human population density and programme phase as significant covariates of programme cost per km^2^ of release area (p = 0.003 and p = 0.026 respectively, two-sided t-test, Appendix page 2). Release material (eggs or adult mosquitoes) or national GDP per capita (as a proxy for local labour costs) were not found to be significant (p = 0.98 and 0.31 respectively) but were retained in the final model due to limited within site variance. Models with the response variable of cost per km^2^ gave superior cost data fit to models with a response variable of cost per person, so were used throughout (Appendix page 2).

Each of the four candidate sites differs in size and human population density, comprising a small city (Yogyakarta City), a large city (Jakarta) and two moderate-size urban-rural mixes (Yogyakarta SAR and Bali, Table 1). Because we assume *Wolbachia* to be suitable only in areas with density greater than 1,000 people per km^2^, only 24.8% and 14.9% of the land area in Yogyakarta SAR and Bali are eligible for *Wolbachia* release, compared to 100% in urban areas, although these areas do still contain the majority of people (Table 1).

**Table 1:**
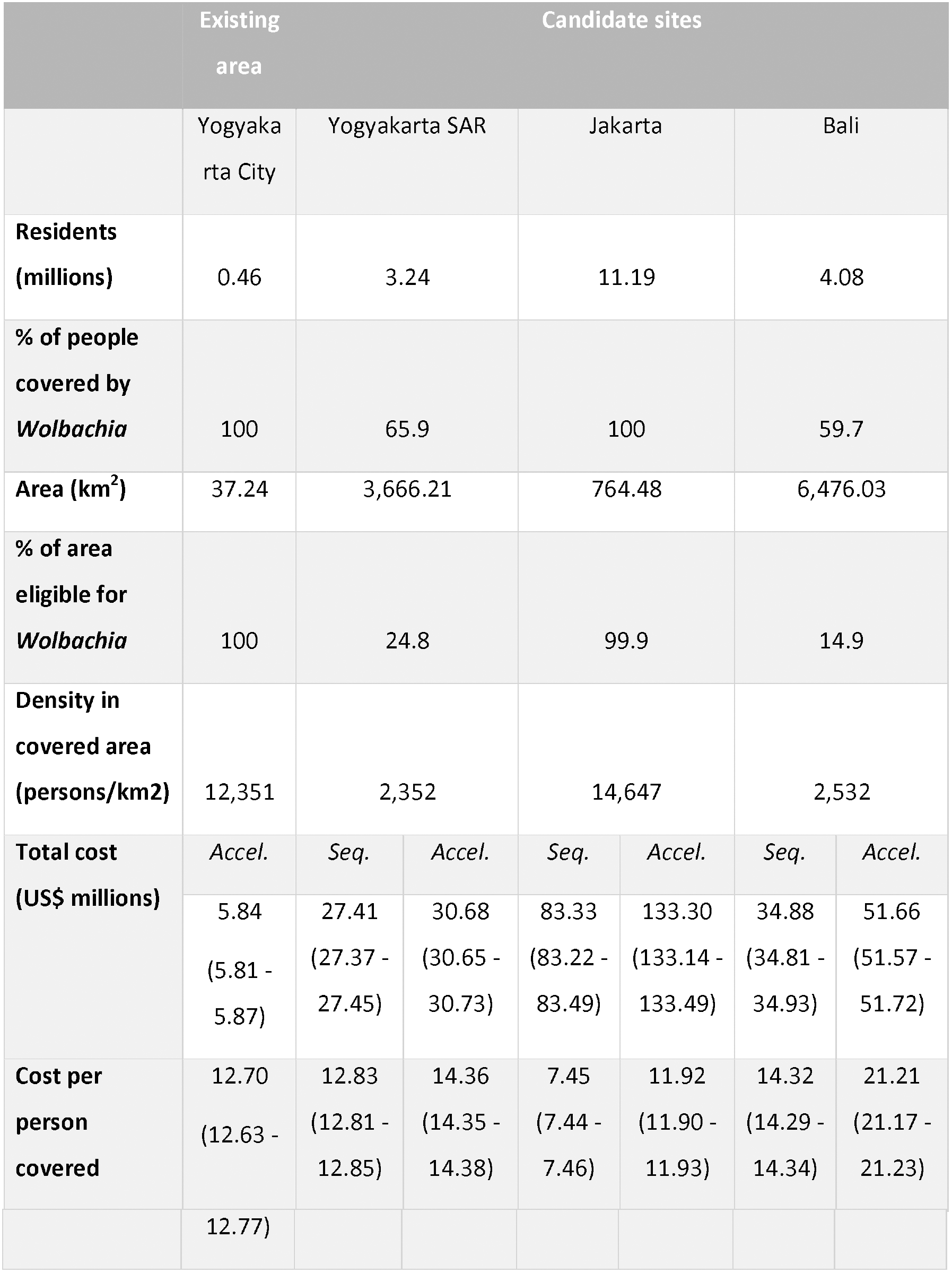
Baseline characteristics and model-predicted release costs for current and future release areas. Prices are in present value 2018 USD. Figures in brackets represent 95% uncertainty intervals. All costs are discounted at 3% per annum. Accel. denotes accelerated; Seq. denotes sequenced.

The estimated cost of an accelerated (10-year) *Wolbachia* programme ranges from $5.8 million in Yogyakarta City to $133.3 million in Jakarta (present value 2018 USD, Table 1). While the urban sites have a smaller release area than their urban-rural mix counterparts, the cost per km^2^ of releasing in high density areas is much higher (Figure 1), however, because more people are covered, urban areas lead to more favourable cost per person covered (∼$12 vs $14-21, Table 1).

Conducting releases over a longer sequenced programme (total programme length 20-year vs 13-year) can reduce overall costs by 11%-38% (Table 1), but also delays benefits (Figure 2). In this analysis we assume 10 years of benefits for each area in which *Wolbachia* mosquitoes are released because there is currently substantial uncertainty over costs and effectiveness beyond 10 years (Figure 2). Should *Wolbachia* prove more durable than this, accelerated programmes and their quicker acquisition of benefits would become more preferable relative to sequenced programmes, however the challenges of their greater upfront costs would remain.

**Figure 2:**
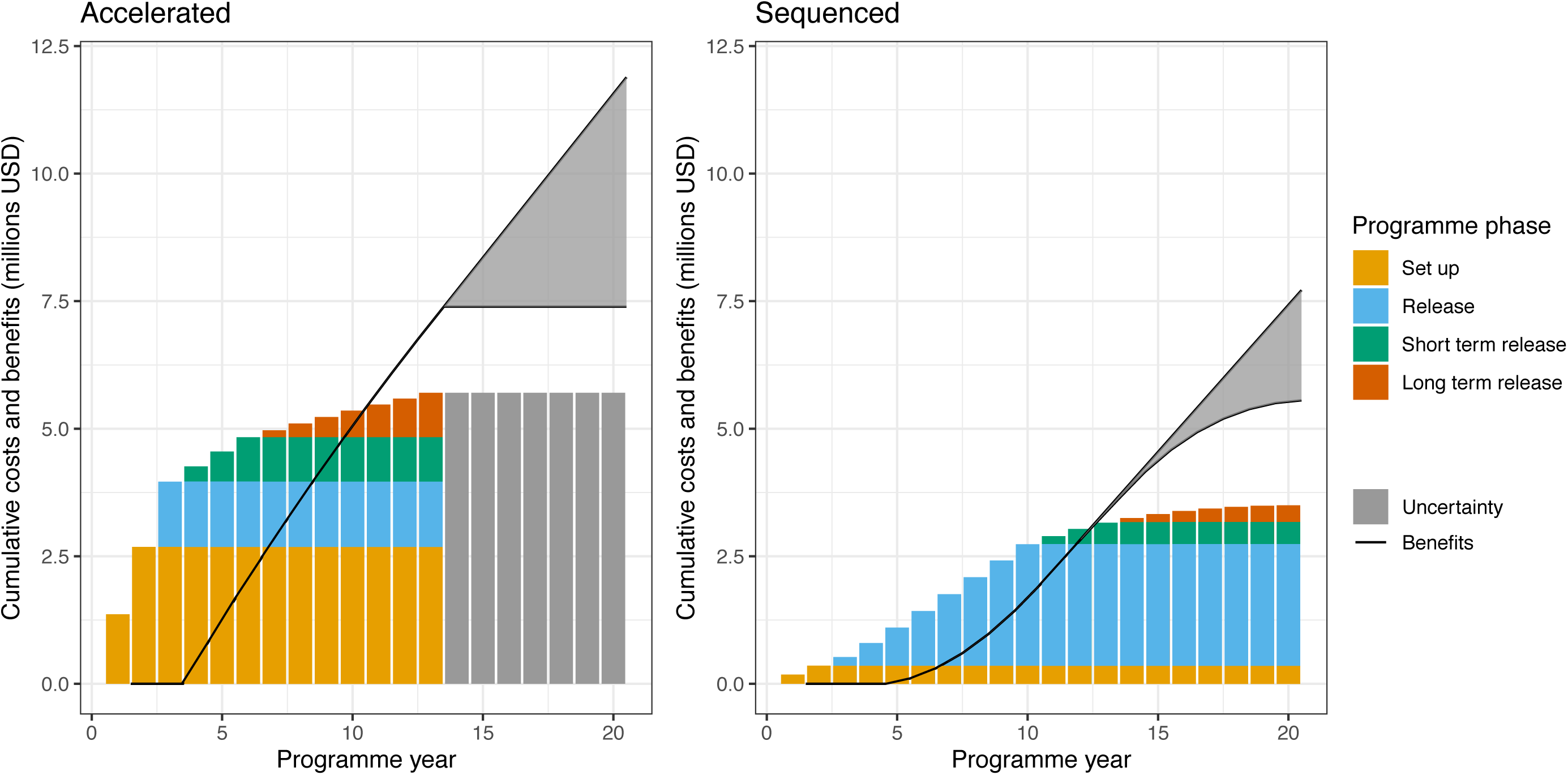
Distribution of cumulative costs and savings over time in an accelerated (3-year, left panel) and sequenced (10-year, right panel) roll out in Yogyakarta City in 2018 USD. Uncertainty represents uncertainty in programme cost and intervention effectiveness beyond the 10-year post-release time horizon used in this analysis. All costs and benefits are in present-day value 2018 USD discounted at 3% per annum.

### Benefits

Combining health systems costs and societal costs (lost wages due to work absences attributable to sickness and the value of life lost due to premature death), Indonesia’s national economic burden of dengue in 2017 has been estimated at $681.26 million, [25] with costs due to hospitalised non-fatal cases (44.7%), fatal cases (44.3%), ambulatory non-fatal cases (5.7%) and non-medical cases (5.3%). [25] We predict substantial reductions in dengue case and economic burden in all sites. As estimated in previous work, [16] long-term average percentage reductions are likely to be highest in low transmission intensity environments (87.2% reduction in Yogyakarta SAR vs 65.7% reduction in Jakarta, Table 2). However, because *Wolbachia* programmes can achieve higher coverage in dense high transmission intensity cities, the percentage reduction across the whole site area becomes more favourable (65.7% in Jakarta vs 59.1% in Yogyakarta SAR and 52.4% in Bali). Medium transmission intensity high density cities, such as Yogyakarta City are likely to see the highest percentage reduction and may even see elimination (94.4%, 95% uncertainty interval [95UI] 36.5%-100%).

**Table 2:** Predicted per year benefits of Wolbachia programmes in four sites. Only eligible areas (at least 1,000 people per km^2^) receive treatment. All costs are in 2018 US dollars and are not discounted. Figures in brackets represent 95% uncertainty intervals.

|  | Committed area | Scale up scenario |  |  |
| --- | --- | --- | --- | --- |
|  | Yogyakarta City | Remaining Yogyakarta SAR | Jakarta | Bali |
| **Baseline cases per year (area wide)** | 14,488<br>(2,955 – 47,858) | 93,604<br>(21,527 – 249,843) | 444,528<br>(97,542 – 1,189,217) | 117,840<br>(27,862 – 324,114) |
| **Post *Wolbachia* cases per year (area wide)** | 722<br>(0 – 30,140) | 36,674<br>(6,805 – 180,567) | 127,712<br>(0 – 749,850) | 62,033<br>(9,810 – 237,984) |
| **Area-wide percent reduction (%)** | 94.4<br>(36.5 – 100) | 59.1<br>(25.6 – 69.6) | 65.7<br>(36.7 – 100) | 52.4<br>(24.2 – 66.9) |
| **Percent reduction within treated areas** | 94.4<br>(36.5 – 100) | 87.2<br>(37.2 – 100) | 65.7<br>(36.7 – 100) | 82.8<br>(37.3 – 100) |
| **Averted medical costs per year (millions USD)\*** | 0.46<br>(0.14 – 1.19) | 2.16<br>(0.67 – 4.70) | 12.91<br>(4.49 – 25.08) | 2.58<br>(0.82 – 5.11) |
| **Averted societal costs per year (millions USD)\*** | 0.52<br>(0.21 – 0.98) | 2.36<br>(1.05 – 4.28) | 14.17<br>(6.09 – 24.27) | 2.77<br>(1.23 – 4.74) |

The annual cost savings of averting these cases are substantial, ranging from $980,000 (95UI $350,000 – $2,170,000) in Yogyakarta City to $27.1 million (95UI $10.58 – $49.35 million) in Jakarta. As estimated in previous work, [25] these cost savings are divided approximately equally between medical costs and societal costs.

### Cost-effectiveness

Due to the heterogeneous nature of risk and cost, estimated cost-effectiveness values are spatially variable (Table 3, Figure 3). Generally, cost-effectiveness is most favourable in high density urban environments with gross cost-effectiveness (cost of averted disease cases not included) reaching as favourable as $1,100 per DALY averted in specific places (Figure 3C), especially in a sequenced programme (Table 3). Although the overall gross cost-effectiveness of the projected programmes in Yogyakarta SAR and Bali are less favourable than their urban counterparts (Table 3), there are many sub-areas within these sites where *Wolbachia* programmes could have equally as favourable cost-effectiveness (Figure 3B and 3D). This is most pronounced for the Yogyakarta SAR scenario where the surrounding urban areas of Sleman, Bantul and the isolated towns of Sentol and Wonosari are predicted to be highly cost effective (<$1,700 / DALY) while many rural areas are less favourable. We even predict some of these towns in Yogyakarta SAR to be more cost effective than Yogyakarta City, however this result occurs only because we assume that the core resources (e.g. laboratory and rearing facilities) that have already been paid for and developed for the existing *Wolbachia* programme in Yogyakarta City can be reused for the surrounding areas in Yogyakarta SAR.

**Table 3:** Predicted cost-effectiveness of Wolbachia at the end of the programme. Accelerated and sequenced programmes correspond to completing roll out in 3 and 10 years, respectively. Only eligible areas (at least 1,000 people per km^2^) receive treatment. All costs are in present value 2018 US dollars. All costs and benefits are discounted at a rate of 3% per annum. Net costs include cost offsets for medical and societal benefits from averted cases. Figures in brackets represent 95% uncertainty intervals.

|  | Committed area | Scale up scenario |  |  |
| --- | --- | --- | --- | --- |
|  | Yogyakarta City | Remaining Yogyakarta SAR | Jakarta | Bali |
|  |  | <u>Accelerated introduction</u> |  |  |
| **Gross \$/DALY averted** | \$ 1,831<br>(892 – 4,522) | \$ 2,133<br>(1,048 – 4,953) | \$ 1,566<br>(857 – 3,244) | \$ 2,996<br>(1,599 – 6,778) |
| **Net \$/DALY averted (including averted medical and societal costs)** | \$ -543<br>(-1,419 – 1,976) | \$ -242<br>(-1,275 – 2,438) | \$ -839<br>(-1,500 – 731) | \$ 671<br>(-719 – 4,219) |
|  |  | <u>Sequenced introduction</u> |  |  |
| **Gross \$/DALY averted** | \$ 1,519<br>(737 – 3,732) | \$ 2,168<br>(1,064 – 5,042) | \$ 1,111<br>(611 – 2,307) | \$ 2,366<br>(1,264 – 5,379) |
| **Net \$/DALY**<br>**averted**<br>**(including health**<br>**sector and societal**<br>**costs)** | **\$ -862**<br>**(-1,572 – 1,185)** | **\$ -210**<br>**(-1,258 – 2,528)** | **\$ -1,280**<br>**(-1,772 – -207)** | **\$ 64**<br>**(-1,050 – 2,834)** |

**Figure 3:**
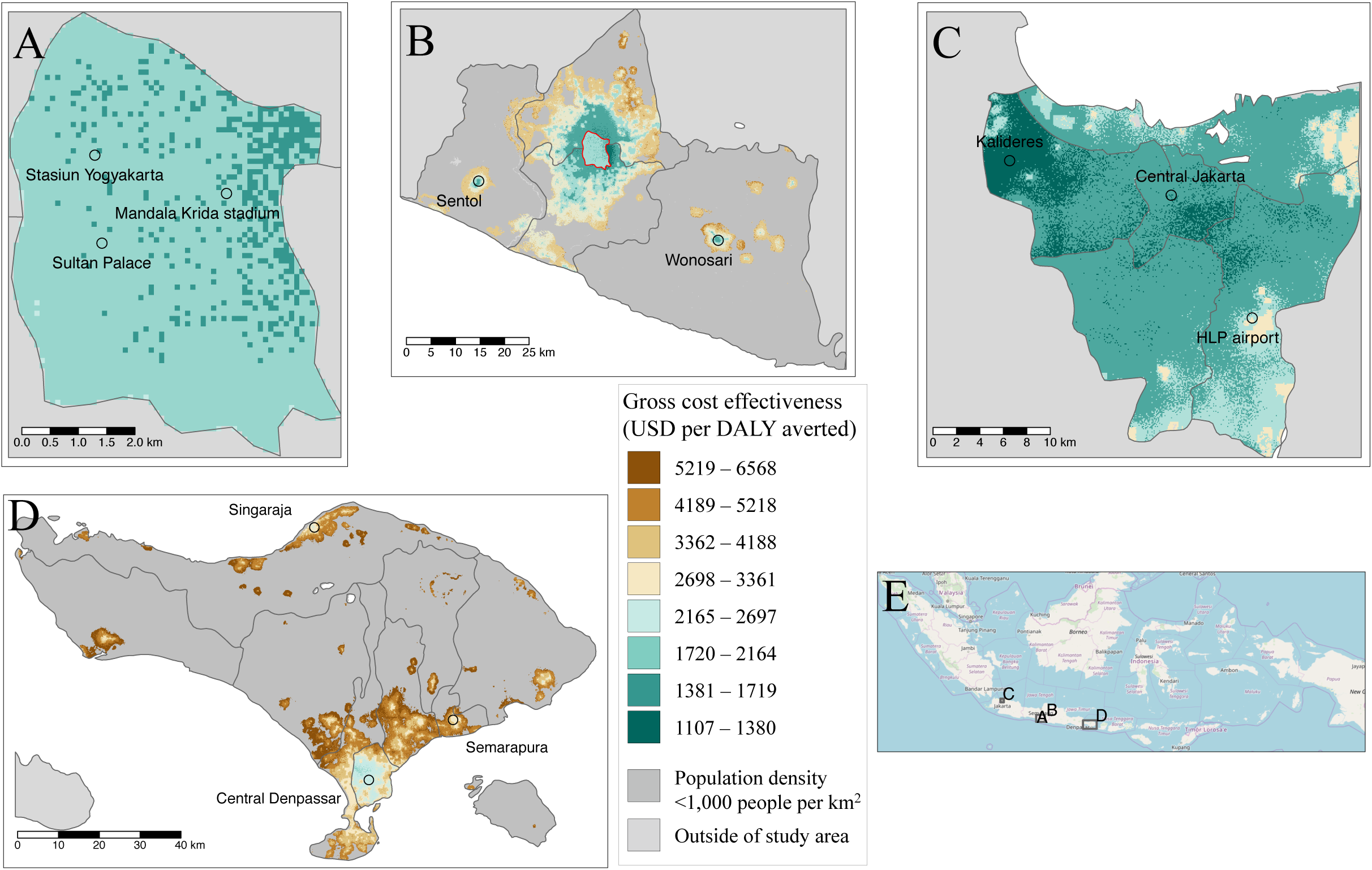
Maps of the gross cost-effectiveness of accelerated Wolbachia releases in Yogyakarta City (A), Yogyakarta SAR (B), Jakarta (C) and Bali (D). Cost effectiveness is measured in present value 2018 USD per Disability Adjusted Life Year (DALY) averted with green areas being most favourable. Select areas of interest and the national orientation of these sites (E) are shown for reference, more detailed background maps are available in the Appendix pages 9-13. Site A falls within site B and is marked in a red outline.

When the health sector and societal costs of averted cases are deducted from the original programme investment, *Wolbachia* becomes a cost-saving intervention in cities and a highly cost-effective intervention elsewhere (Table 3 and Figure 4A). One dollar invested in *Wolbachia* can return between $1.35 and $3.40 (95UI $0.17 – $9.67) in medical and societal benefits depending on where the programme takes place (Figure 4A). In Jakarta, the medical benefits alone are predicted to outweigh the cost of investment in *Wolbachia* (Figure 4A).

**Figure 4:**
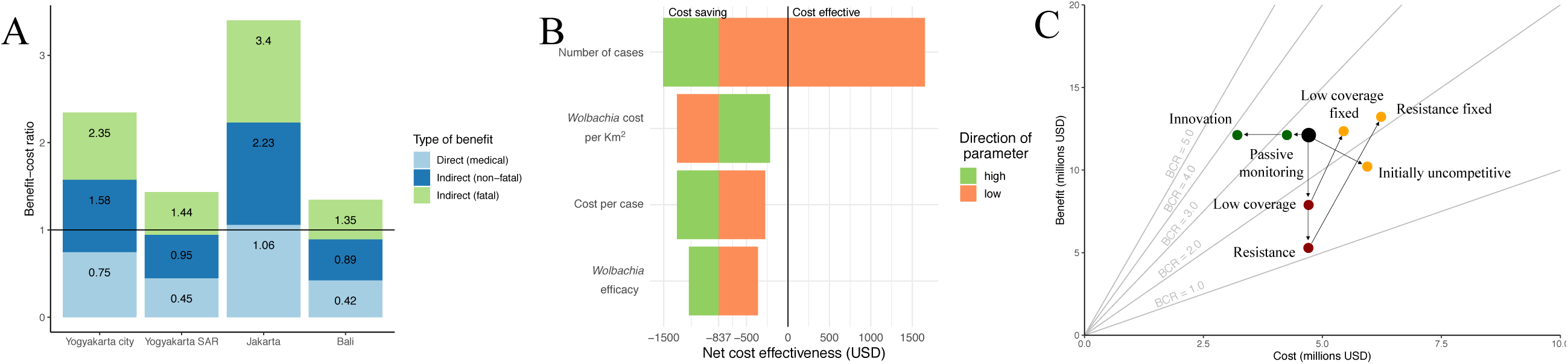
Benefit-cost ratios and their sensitivity. 4A) The predicted benefit-cost ratios of a sequenced release programme in each site disaggregated by type of benefit. A benefit-cost ratio of 1.0 indicates cost saving. 4B) Sensitivity of cost-effectiveness of a sequenced release in Yogyakarta City if the 2.5% value (orange) or 97.5% value (green) were used as opposed to the median value for selected parameters. Net cost-effectiveness is in 2018 present day value USD per Disability Adjusted Life Year averted and includes offsets from health sector and societal perspectives. 4C) Sensitivity of benefit-cost ratios (BCRs) to future challenges or changes to a sequenced release in Yogyakarta City. Green dots show potential cost-saving measures, red dots unaddressed challenges to the programme and yellow dots addressed challenges to the programme. Dots above the BCR = 1.0 line are cost saving from the societal perspective.

#### Sensitivity and uncertainty

Our prediction that *Wolbachia* is cost saving is robust to a reasonable range of parameter values (Figure 4B). In univariate sensitivity analysis of the 2.5 and 97.5 percentiles of the true parameter value, only a low value of the baseline burden of dengue is sufficient to prevent *Wolbachia* from becoming cost saving, and even then the programme is still highly cost effective ($1652 / DALY, Sequenced programme in Yogyakarta City, societal perspective). Parameters for cost of the programme, cost per case and efficacy of the intervention were less critical for overall cost-effectiveness than baseline burden due to the higher uncertainty in the true burden of dengue.

Programmes are even predicted to be cost saving if substantial challenges occur during deployment. If only 50% coverage were reached in the target area, resistance emerges after five years or released mosquitoes are uncompetitive with wild-type mosquitoes, benefit-cost ratios remain above 1 (Figure 4C, societal perspective). Furthermore, if these events do occur, cost-effectiveness of the programme can still be recovered by addressing these threats (Figure 4C and Appendix page 4-6). As *Wolbachia* programmes become more proven over time it is expected that relaxed surveillance (relying on passive disease monitoring), innovations and economies of scale will reduce the cost of deployment. These have the potential to increase the benefit-cost ratio by as much as 47%, as long as they do not come at the expense of avoiding to detect more damaging failures of the programme (Figure 4C).

## Discussion

Here we use existing cost data to build a programmatic model for *wMel Wolbachia*. By applying this model in Indonesia we show that this novel technology can be an economically advantageous intervention for dengue control and predict under what circumstances it might be most cost effective. Cost effectiveness of *Wolbachia* is predicted to be most favourable in dense cities where a high concentration of people and dengue incidence allow the high investment costs of *Wolbachia* to be quickly offset. In such areas, programmes can become cost saving with benefit to cost ratios of 1.35-3.40, which are robust to our choice of model parameters and difficulties in the programme. Finally, we show that *Wolbachia* can also be cost effective in suburban and rural areas, particularly if they can utilise programme infrastructure from nearby urban areas. This is particularly relevant for the existing *Wolbachia* programme in Yogyakarta City and suggests that expansion to nearby areas in Sleman and Bantul in Yogyakarta SAR should be considered.

Assessing the cost-effectiveness of novel rear and release vector control strategies is important because of their high upfront costs but potentially long-term benefits. This makes the cost-effectiveness dynamics of *wMel Wolbachia* more similar to mass vaccination than conventional vector control. Like vaccination, this makes cost-effectiveness of *Wolbachia* sensitive to the time horizon of the evaluation. *wMel Wolbachia* has been robustly established in Cairns, Australia since early 2011 [26], hence our assumed 10 year benefit time horizon. More research is required to understand the sustainability of *Wolbachia* replacement in dengue-endemic countries with more complex *Aedes* population genetics and higher virus and mosquito importation rates from outside areas [30].

A number of previous studies have attempted to estimate the cost-effectiveness of vector control interventions for dengue [20, 31–34]. The methods used tend to fall into one of two approaches: field trials or model-based assessments. Experimental and observational control trials have been used to estimate cost-effectiveness (per DALY averted) for larvicides in Cambodia ($313) [31], community clear up campaigns ($3,953) and ultra-low volume spraying ($4,472) in Mexico [33] and an integrated package of vector control interventions in Sri Lanka ($98) [34]. Short-term control trials with disease endpoints are likely to overestimate effectiveness due to the effects of heard immunity and may mean interventions delay rather than avert disease.

In response to this, model-based cost-effectiveness evaluations can be used to give a more accurate estimate of long-term effectiveness of a particular intervention. However, because long-term effectiveness is not easily measurable, such modelling studies often have to assume a range of plausible efficacies with variable theoretical support. Modelling studies have suggested larval control ($615-1,267 / DALY) [32] and more generic packages of vector control ($679-1907 / DALY) [20] can also be cost effective depending on true effectiveness. Finally, several models have predicted the cost-effectiveness of dengue vaccines [22, 35]. The cost-effectiveness of WHO’s recommended test-and-vaccinate strategy in Indonesia in 2015 was 0.8 to 0.6 times the per capita GDP (i.e., $2,700 and $2,000) if dengue seroprevalence rates at age 9 were 50% and 70%, respectively [22]. Despite vaccination having a less favourable predicted cost-effectiveness than *Wolbachia*, it is likely both vaccine and vector control will be necessary to achieve control in the highest transmission areas. Further work is needed to understand how the economics of combinations of interventions vary across transmission strata.

Given that *Wolbachia* is also not predicted to fully eliminate dengue virus transmission in highly endemic settings [16, 17] and given that many countries already have established dengue control programmes, there is a pressing need to understand how *Wolbachia* interacts with other types of vector control and how the optimal package of interventions may change in different environments. Modelling and mapping techniques are critical for such investigations due to the impracticality of conducting field trials among the many combinations of different interventions [36, 37].

Our approach to assessing the cost-effectiveness of *Wolbachia* combines the best currently available evidence for the effect of *Wolbachia* on transmission [38] with a long-term mathematical model [16] to overcome limitations of both of these approaches. This work aims to provide an evidence-based first estimate that gives quantitative support behind the decision to invest large sums of money in an intervention that is likely to have deferred but substantial benefits. Using model-based estimates of the true case and economic burden of dengue [16, 25] in Indonesia was a critical step in our approach. Using reported case data would have significantly underestimated the cost-effectiveness of *Wolbachia* and more research is needed to understand, adjust for and ultimately fill gaps in disease surveillance [39].

This analysis was subject to a number of limitations. First, our model did not consider logistical constraints that may exist in releasing *Wolbachia* infected mosquitoes at this scale. The largest current planned releases of *Wolbachia* mosquitoes is in Medellin Colombia where a sequenced programme will cover a combined 1.7 million people over 151km^2^. Reaching high coverage of *Wolbachia* for Jakarta’s 11 million residents and 764 km^2^ land area, particularly over a three-year accelerated campaign, may not be logistically feasible. New approaches to large scale community engagement and recruitment of release teams need to be developed. There may also be constraints on the portability of assets, such as centralised distribution of mosquitoes or laboratory testing, across areas as wide as Bali that we did not consider. Second, cost data for existing *Wolbachia* releases were based on budgeted costs; actual costs may differ by the end of the programme. Third, our analysis only included the effects of *Wolbachia* on dengue, despite showing strong protective effects against a range of other arboviral diseases [3–5]. Given chikungunya is also ubiquitous in Indonesia [40] our predictions may underestimate the cost-effectiveness of *Wolbachia*. Finally, it should be mentioned that the cost-effectiveness analysis presented here is intended to form one part of the wider evidence base on whether or where *Wolbachia* should be scaled up. To date, successful *Wolbachia* programmes have been underpinned by sustained and robust engagement with both the community and local stakeholders [13, 41]. In this analysis we make clear assumptions about the success of establishing *Wolbachia* in a target area, but clearly an assessment of feasibility of this aim is a necessary precursor to assessments of cost-effectiveness.

The biggest strength of our analysis is the use of comprehensive, detailed spatiotemporal models that incorporate the latest projections of dengue case and economic burden and the likely impact *Wolbachia* could have on when deployed at scale. Given *Wolbachia* is an early stage novel intervention, we have also endeavoured to include the broad range of uncertainty that exists in each of these inputs and assess their impact overall cost-effectiveness. Such comparisons are important if the high upfront investment costs of *Wolbachia* are to be justified and these results can be used as part of the evidence base in the decision to accelerate scale up of *Wolbachia* to address the growing needs of arboviral control.

## Conclusions

In conclusion, in this study we show that *Wolbachia* has the potential to be a highly cost effective and even cost saving intervention, especially if targeted to high density cities where the burden of dengue is concentrated. These findings are largely robust to uncertainties in the long-term performance of *Wolbachia*, but further longitudinal field data with epidemiological outcome measures are required to validate these predictions and assess how cost-effectiveness changes when combined with other vector control interventions and vaccines.

## Data Availability

All data used in this analysis is publicly available and can be accessed in the related paper O’Reilly et al. BMC Medicine 2019 17:172 or in the Appendix of this paper. All code to reproduce our analysis is publicly available in the following GitHub repository: (https://github.com/obrady/Wolbachia_CE/tree/V1).

## List of abbreviations

CEA: Cost Effectiveness Analysis DALY Disability Adjusted Life Year
GDP: Gross Domestic Product PPP Purchasing Power Parity
SAR: Special Autonomous Region
USD: United States Dollar
WHO: World Health Organization
WMP: World Mosquito Programme

## Declarations

### Ethics approval and consent to participate

Not applicable

### Consent for publication

Not applicable

### Availability of data and materials

All data used in this analysis is publicly available and can be accessed in the related paper O’Reilly et al. [16] or in the Appendix of this paper. All code to reproduce our analysis is publicly available in the following GitHub repository: (https://github.com/obrady/Wolbachia_CE/tree/V1).

### Authors’ contributions

OJB, DSS conceived and designed the study. KMO, OJB, EH, DDK, NNW and LSB analysed the data. OJB, DDK, NNW, KMO, EH, LSB, LY and DSS drafted and revised the manuscript. All authors read and approved the final manuscript.

## Acknowledgements

The design of the study was developed independently, but benefitted from a series of in-depth discussions with representatives from WMP global headquarters in Melbourne Australia (including Cameron Simmons, Scott O’Neil and Reynold Dias) and field site coordinators from UGM, Yogyakarta city, Indonesia (including Adi Uterini). The investigators had access to all the data and had final responsibility for the decision to submit for publication.

